# Pharmacoepidemiology of Cardiovascular Medicines Used Among Inpatients at a Tertiary Care Hospital in Tanzania, 2016-2022

**DOI:** 10.64898/2025.11.30.25341284

**Authors:** Philip Sasi, Justine L. Vasco, Raphael Z. Sangeda, Tusaligwe Mbilinyi, Naizihijwa Majani, Peter Richard Kisenge, Pilly Chillo

## Abstract

**Introduction:** Cardiovascular diseases remain a major cause of morbidity and mortality in sub-Saharan Africa; however, little is known about inpatient cardiovascular medicine utilisation in Tanzania. This study quantified cardiovascular medicine utilisation patterns at a national cardiac referral centre using the standardised WHO ATC/DDD methodology.

**Methods:** A retrospective longitudinal pharmacoepidemiological study was conducted using inpatient prescription data from the Jakaya Kikwete Cardiac Institute (JKCI) for the fiscal years 2016/2017 to 2021/2022. Non-medicinal items and topical formulations were excluded from the analyses. Utilisation was expressed as Defined Daily Doses (DDD) per 100 bed-days using the WHO ATC/DDD methodology. Descriptive statistics and non-parametric tests were performed using SPSS version 26.

**Results:** Of the 290,591 prescription records screened, 224,290 were eligible following the exclusion of 64,781 non-medicinal items and 1,520 topical agents. Cardiovascular medicines (ATC class C) accounted for 99,634 (44.4%) of all prescriptions, followed by alimentary tract and metabolism agents (18.1%). The prescribing volumes increased annually and peaked in 2021/2022.

Furosemide (C03CA01), spironolactone (C03DA01) and paracetamol (N02BE01) were the three most frequently prescribed medicines. The 12 most commonly used medicines represented approximately half of all the prescriptions.

**Conclusion:** Cardiovascular medicines accounted for nearly half of all inpatient prescriptions, reflecting the substantial clinical demand and disease burden associated with cardiovascular conditions.

Utilisation was dominated by loop and potassium-sparing diuretics, highlighting their central role in inpatient cardiac care. Routine surveillance using ATC/DDD metrics should be institutionalised to guide procurement, prescribing audits, and patient safety monitoring.

Trial registration number

Not applicable.

*What is already known on this topic:*

- The incidence of cardiovascular diseases is increasing globally, with increasing medicine use in both high-and low-income settings.
- Evidence of inpatient cardiovascular medicine consumption in sub-Saharan Africa is limited to a few studies.
- Previous pharmacoepidemiological studies from SSA have mainly focused on antibiotics rather than medicines for chronic noncommunicable diseases.

*What this study adds”:*

- This is the first longitudinal analysis of inpatient cardiovascular medicine use in Tanzania based on ATC/DDD.
- Nearly half of all inpatient prescriptions at the national cardiac referral centre were cardiovascular agents, predominantly diuretics.
- These findings establish a baseline for stewardship, informed procurement decisions, and future national monitoring efforts.

## Introductions

Cardiovascular diseases (CVDs), including coronary heart disease, cerebrovascular disease, peripheral arterial disease, rheumatic and congenital heart disease, deep-vein thrombosis and pulmonary embolism, are the leading causes of morbidity and mortality globally [1–6]. The burden is driven by both traditional and non-traditional risk factors, including hypertension, diabetes, obesity, physical inactivity, tobacco and harmful alcohol use, and air pollution [7–11]. Women, especially postmenopausal women, experience additional risks linked to hormonal and metabolic transitions [8–10]. Familial predisposition, childhood obesity and chronic metabolic conditions further amplify lifetime CVD vulnerability [11–13].

Pharmacoepidemiology, which links pharmacology and epidemiology, quantifies medicine utilisation in real-world populations [12] and is particularly relevant in CVDs, where multi-drug therapy is the norm. It enables the measurement of trends, exposure intensity, and rational-use indicators and supports stewardship, safety surveillance, and resource planning.

The WHO Anatomical Therapeutic Chemical (ATC) classification system and Defined Daily Dose (DDD) methodology are international standards for quantifying medicine consumption and for cross-institutional comparisons [14]. However, despite the increasing CVD burden in sub-Saharan Africa, inpatient cardiovascular medicine utilisation patterns in Tanzania remain poorly understood.

Despite the growing burden of cardiovascular disease in Tanzania, limited data exist on inpatient prescribing patterns in tertiary cardiac centres. The Jakaya Kikwete Cardiac Institute (JKCI), Tanzania’s national tertiary cardiac referral centre, provides an opportunity to address this knowledge gap. Therefore, this study aimed to describe the prescribing patterns and consumption trends of cardiovascular medicines among inpatients at JKCI between 2016 and 2022 using the WHO ATC/DDD methodology.

## Methods

### Study design and setting

This was a retrospective longitudinal pharmacoepidemiological study of cardiovascular medicine utilisation among inpatients at the Jakaya Kikwete Cardiac Institute (JKCI), Dar es Salaam, Tanzania, covering the fiscal years 2016/2017 through 2021/2022. JKCI is the national cardiac referral centre (approximately 150 beds) and an academic affiliate of Muhimbili University of Health and Allied Sciences (MUHAS).

### Study population and data source

All inpatients admitted to the JKCI during the study period who received cardiovascular medicines (ATC level 1, Class C) were included. Dispensing data were obtained from *MedPro*, a JKCI electronic pharmacy information system that captures patient-level demographic and prescription data. The extracted variables included sex, age group, sponsor type, admission date, ATC code, dosage form, strength, quantity dispensed, and route of administration.

### Inclusion and exclusion criteria

All systemic cardiovascular medicines (ATC level 1 Class C) and co-prescribed systemic antibiotics were included to account for procedure-related prophylaxis and therapeutic exposures. Topical, ophthalmic, otic, and dermatologic preparations were excluded to maintain consistency in systemic exposure measurements and ensure quantification using the World Health Organisation Anatomical Therapeutic Chemical/Defined Daily Dose (WHO ATC/DDD) methodology.

### ATC/DDD standardisation

Medicine utilisation was quantified using the WHO ATC/DDD methodology. For inpatients, the primary outcome metric was Defined Daily Doses per 100 bed-days (DDD/100 bed-days), as recommended for hospital-level monitoring. The total DDDs dispensed were mapped to the WHO ATC Level-5 generic codes and their corresponding WHO standard DDD values.

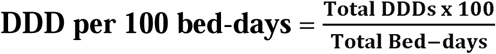

In addition, because the JKCI maintains detailed operational metrics (bed capacity and service-day coverage), we performed extended denominator normalisation. This allows for adjustments not only for occupancy but also for variability in the number of inpatient service days within a fiscal cycle.

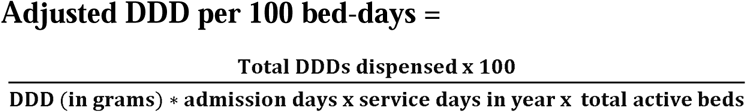

The WHO DDD Index was used for all ATC–DDD values. For interpretive clarity, the DDD values reported in the text and tables were rounded to two decimal places, whereas all underlying calculations retained full precision.

### Drug utilisation concentration analysis

Drug utilisation (DU) concentration was assessed using the DU method. First, all dispensed medicines across ATC level 1 classes (A–V) were ranked by descending dispensing frequency to describe the overall utilisation concentration at the hospital level. Subsequently, cardiovascular agents (ATC level 1, Class C) were isolated, and DU analysis was repeated using the cumulative DDD contribution. DU50 was defined as the group of medicines accounting for the first 50% of cumulative utilisation, and DU90 as the group accounting for the first 90% of dispensing frequency volumes. For ATC level 1 class C medicines, the DU50 and DU90 classifications were based on the cumulative DDD volume (expressed as DDD per 100 bed days). For all medicines (A–V), the DU50 and DU90 classifications were based on dispensing frequency ranking. Inpatient antibiotic DDD analyses from the same source population have been previously reported [15].

## Statistical analysis

Following preprocessing, the datasets were imported into SPSS (version 26). Descriptive statistics were used to summarise patient characteristics and annual utilisation. The normality of the DDD distributions was tested using the Kolmogorov–Smirnov test, which confirmed non-normality. Group-wise differences in DDD per 100 bed days across demographic and sponsor categories were evaluated using Kruskal-Wallis tests, with post-hoc pairwise comparisons where appropriate.

Time-series analysis was used to assess monotonic trends using the Mann–Kendall test. Seasonality was evaluated by visual inspection of monthly means and Kruskal–Wallis testing of month-to-month variation. As a sensitivity analysis, an exploratory study using AutoRegressive Integrated Moving Average (ARIMA) modelling (forecast package in R) examined the longitudinal trend structure, and the Akaike Information Criterion guided model selection. For interpretive clarity, the DDD values reported in the main text were rounded to two decimal places, whereas the underlying computations retained full precision.

## Ethical considerations

This retrospective analysis of de-identified inpatient dispensing data was approved by the Muhimbili University of Health and Allied Sciences Research Ethics Committee (Ref: DA.25/111/01B/184). The requirement for informed consent was waived because the dataset contained no direct personal identifiers. Data processing steps (Power Query transformation, ATC-DDD mapping, and removal of personal identifiers) were performed in a secure JKCI environment prior to export.

## Results

### Overall utilisation of all medicine classes at JKCI

A total of 290,591 inpatient prescription records were retrieved for the 2016/2017–2021/2022 fiscal year. Of these, 66,301 (22.8%) were excluded: 64,781 (22.3%) were non-medical entries and 1,520 (0.5%) were topical external preparations. The remaining 224,290 (77.2%) eligible inpatient prescription records met the inclusion criteria and were retained for analysis (Table 1). Within this analytic dataset, 99,634 (44.4%) prescriptions were for cardiovascular medicines (ATC Class C), whereas 124,656 (55.6%) were prescriptions for other systemic ATC classes.

**Table 1:** Quantity of medicine prescriptions issued to inpatients attending JKCI from 2016/17 to 2021/22.

| **Fiscal Year** | **Number of prescriptions** | **%** |
| --- | --- | --- |
| 2016-2017 | 2,621 | 1.2 |
| 2017-2018 | 29,826 | 13.3 |
| 2018-2019 | 42,769 | 19.1 |
| 2019-2020 | 45,616 | 20.3 |
| 2020-2021 | 49,852 | 22.2 |
| 2021-2022 | 53,606 | 23.9 |
| **Total** | **224,290** | 100 |

These prescription records corresponded to 11,656 unique inpatients over six years.

A subset of prescriptions was cardiovascular medicines (ATC Class C), which accounted for 99,634 (44.4%) of all eligible inpatient prescriptions. This was followed by alimentary tract and metabolism medicines (40,634, 18.1%) and systemic anti-infectives (31,594, 14.1%) (Table 2). The prescription volume has increased progressively over the past few fiscal years, with the highest counts recorded in the 2021/2022 fiscal year (Table 2).

**Table 2:**
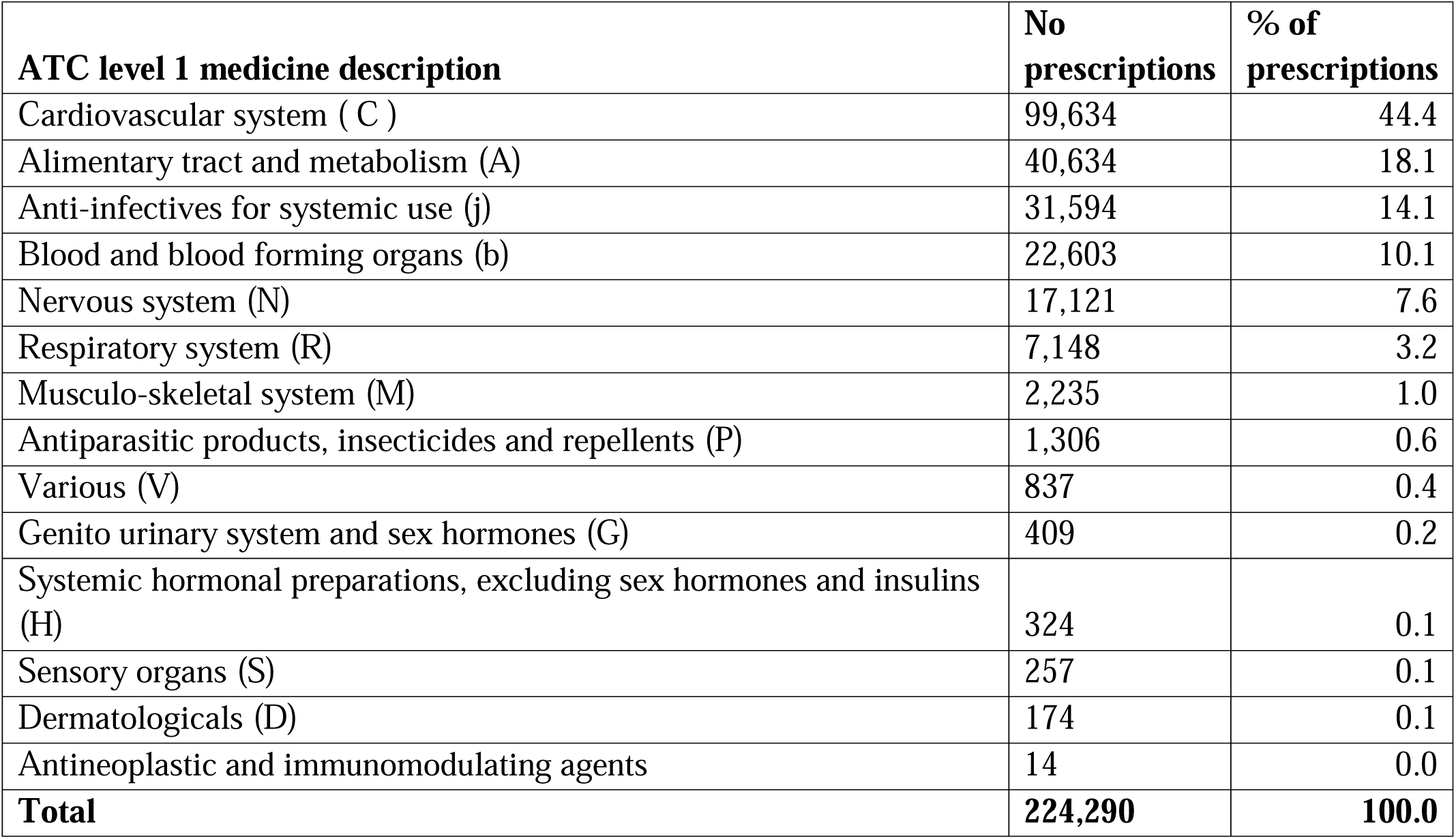
Number of prescriptions per ATC class level 1 medicines.

| **ATC level 1 medicine description** | **No prescriptions** | **% of prescriptions** |
| --- | --- | --- |
| Cardiovascular system ( C ) | 99,634 | 44.4 |
| Alimentary tract and metabolism (A) | 40,634 | 18.1 |
| Anti-infectives for systemic use (j) | 31,594 | 14.1 |
| Blood and blood forming organs (b) | 22,603 | 10.1 |
| Nervous system (N) | 17,121 | 7.6 |
| Respiratory system (R) | 7,148 | 3.2 |
| Musculo-skeletal system (M) | 2,235 | 1.0 |
| Antiparasitic products, insecticides and repellents (P) | 1,306 | 0.6 |
| Various (V) | 837 | 0.4 |
| Genito urinary system and sex hormones (G) | 409 | 0.2 |
| Systemic hormonal preparations, excluding sex hormones and insulins (H) | 324 | 0.1 |
| Sensory organs (S) | 257 | 0.1 |
| Dermatologicals (D) | 174 | 0.1 |
| Antineoplastic and immunomodulating agents | 14 | 0.0 |
| **Total** | **224,290** | **100.0** |

The number of prescriptions for cardiovascular medicines (ATC Class C) was highest in 2021/2022 compared with all other ATC classes in the same fiscal year, followed by medicines in ATC Class A (Figure 1).

**Figure 1:**
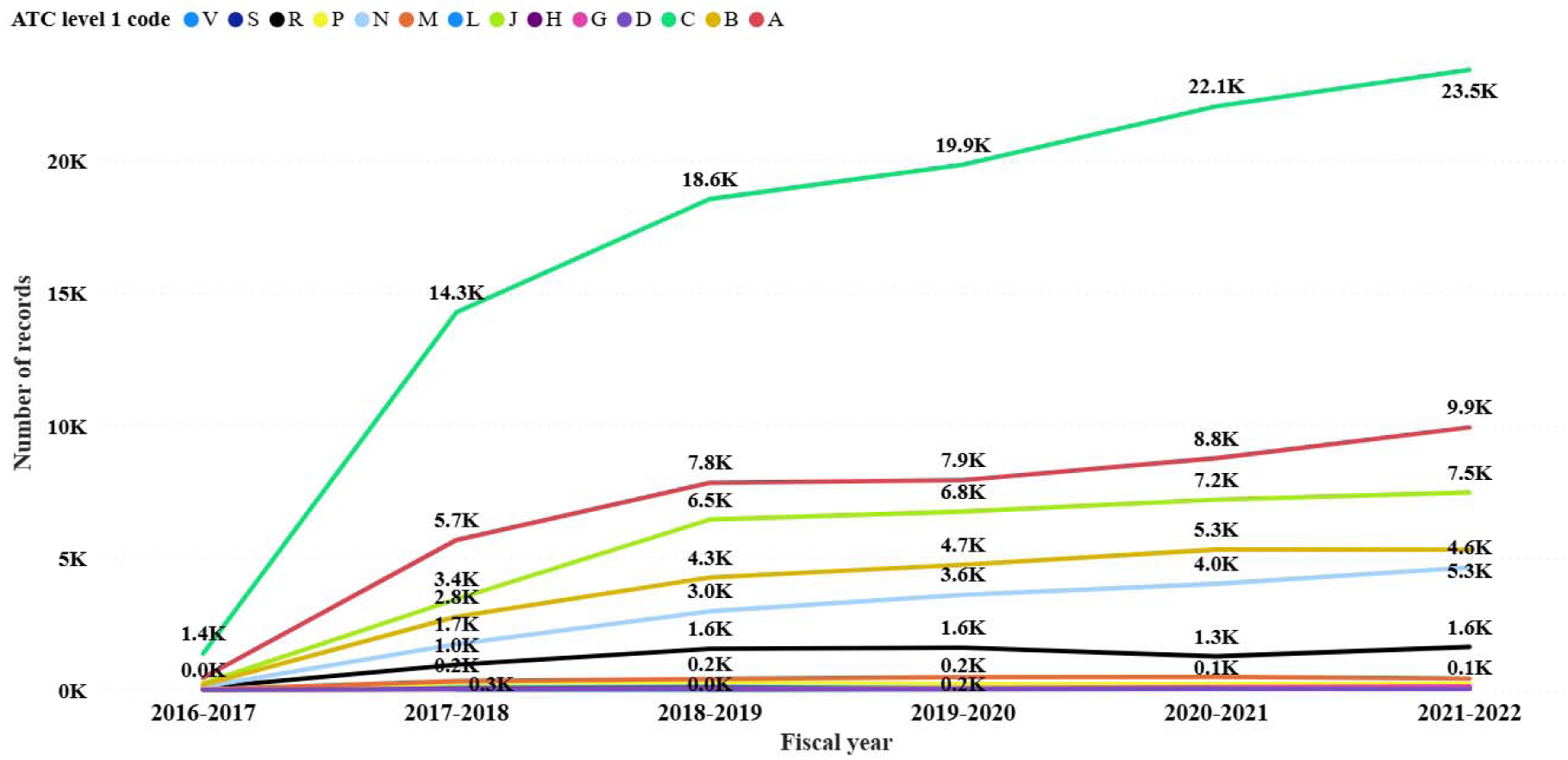
Distribution of prescription counts across ATC Level-1 classes over the study period (2016/2017–2021/2022) for medicines used at the Jakaya Kikwete Cardiac Institute (JKCI).

Key for ATC codes level 1: A = Alimentary Tract and Metabolism; B = Blood and Blood Forming Organs; C = Cardiovascular System; D = Dermatologicals; G = Genito-urinary System and Sex Hormones; H = Systemic Hormonal Preparations Excluding. Sex hormones and insulin; J = anti-infectives for systemic use; L = antineoplastic and immunomodulating agents; M = musculoskeletal system; N = nervous system; P = antiparasitic products, insecticides, and repellents; R = respiratory system; S = sensory organs; and V = various.

The top 12 ATC level 5 medicines accounted for approximately 50% of all inpatient prescription records, representing the DU50 frequency segment at the hospital level (Table 3). Furosemide (C03CA01) was the most frequently dispensed medicine, followed by spironolactone (C03DA01) and paracetamol (N02BE01). In total, 255 unique medicines were used at JKCI (Supplementary Table 1).

**Table 3:**
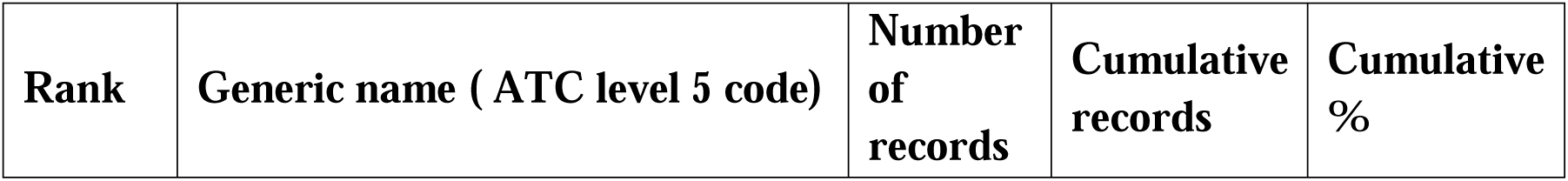
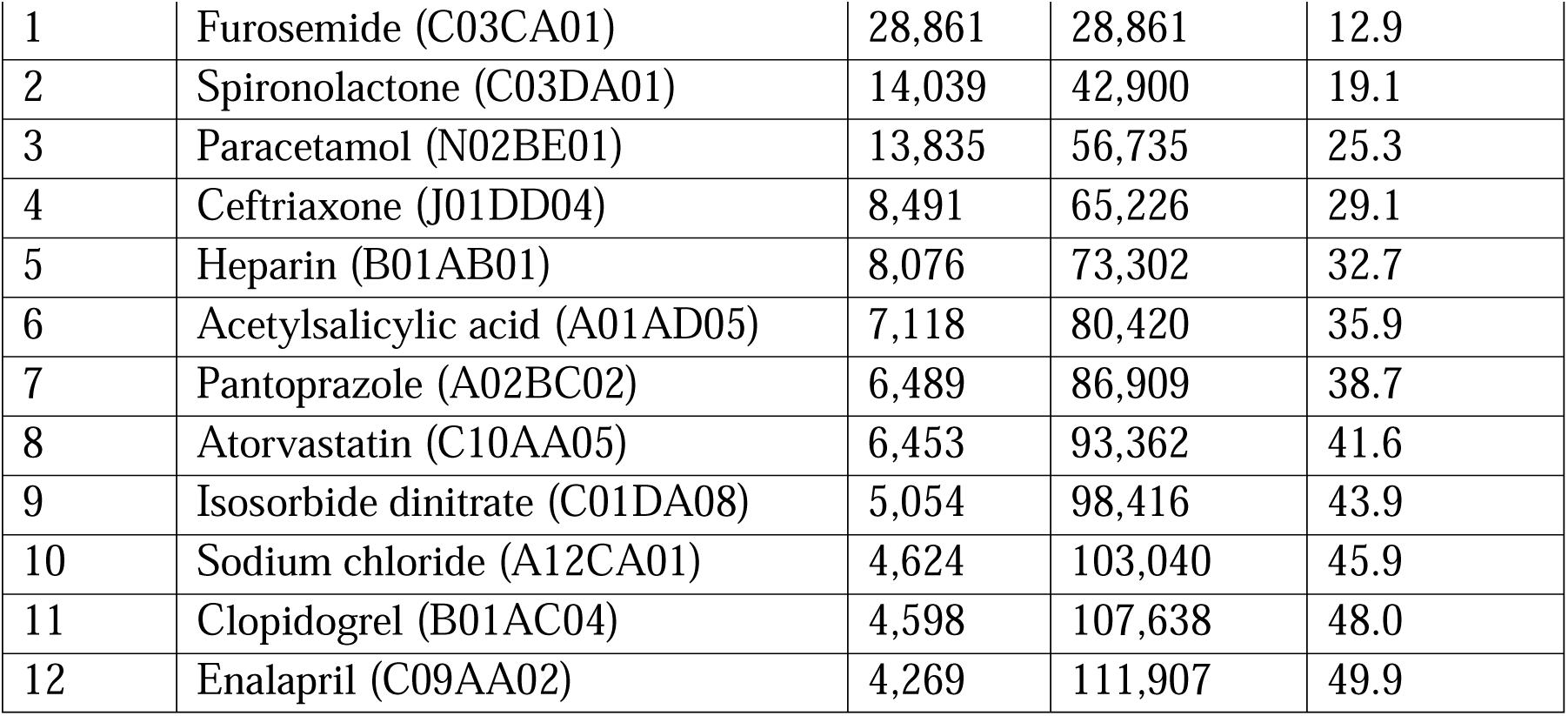
Composition of prescription records distribution for the top 12 ATC level 5 medicines utilised.

### Characteristics of cardiovascular medicine recipients

Of the 11,656 unique inpatients who received one or more medicines during the six-year study period, 9,902 (84.9%) received one or more cardiovascular medicines (ATC Class C). This formed the analytic population for the utilisation intensity analyses (Table 4). Among the 9,902 users of cardiovascular medicine, 5,895 were female (59.5%) and 4,007 were male (40.5%).

**Table 4.**
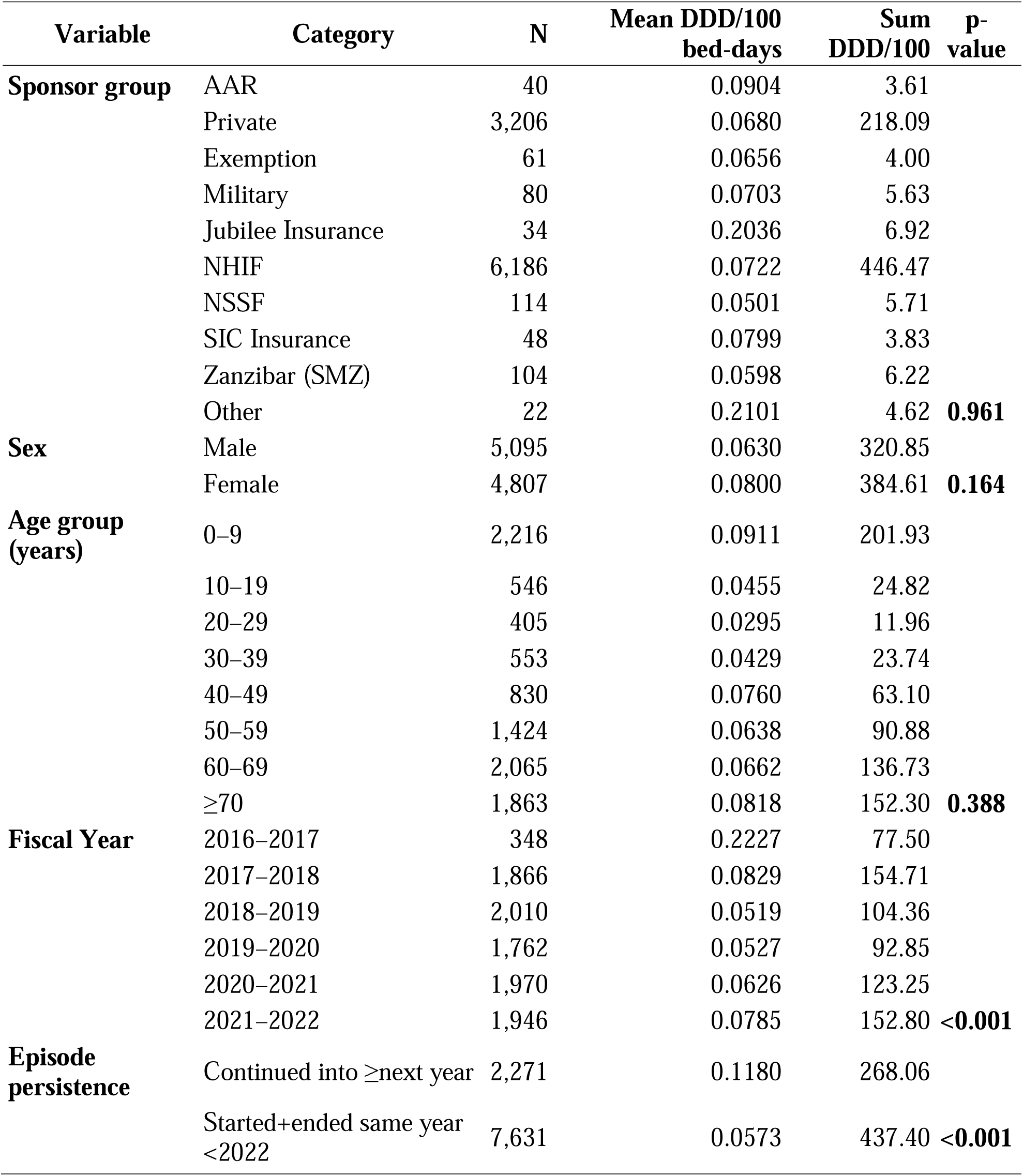
Characteristics of inpatients who received. ≥**1 cardiovascular medicine (ATC Class C) and utilisation intensity (DDD per 100 bed-days) across subgroups at the Jakaya Kikwete Cardiac Institute (JKCI) (N = 9,902), fiscal years 2016/2017–2021/2022.**

| Variable | Category | N | Mean DDD/100<br>bed-days | Sum<br>DDD/100 | p-<br>value |
| --- | --- | --- | --- | --- | --- |
| **Sponsor group** | AAR | 40 | 0.0904 | 3.61 |  |
|  | Private | 3,206 | 0.0680 | 218.09 |  |
|  | Exemption | 61 | 0.0656 | 4.00 |  |
|  | Military | 80 | 0.0703 | 5.63 |  |
|  | Jubilee Insurance | 34 | 0.2036 | 6.92 |  |
|  | NHIF | 6,186 | 0.0722 | 446.47 |  |
|  | NSSF | 114 | 0.0501 | 5.71 |  |
|  | SIC Insurance | 48 | 0.0799 | 3.83 |  |
|  | Zanzibar (SMZ) | 104 | 0.0598 | 6.22 |  |
|  | Other | 22 | 0.2101 | 4.62 | **0.961** |
| **Sex** | Male | 5,095 | 0.0630 | 320.85 |  |
|  | Female | 4,807 | 0.0800 | 384.61 | **0.164** |
| **Age group**<br>**(years)** | 0–9 | 2,216 | 0.0911 | 201.93 |  |
|  | 10–19 | 546 | 0.0455 | 24.82 |  |
|  | 20–29 | 405 | 0.0295 | 11.96 |  |
|  | 30–39 | 553 | 0.0429 | 23.74 |  |
|  | 40–49 | 830 | 0.0760 | 63.10 |  |
|  | 50–59 | 1,424 | 0.0638 | 90.88 |  |
|  | 60–69 | 2,065 | 0.0662 | 136.73 |  |
|  | ≥70 | 1,863 | 0.0818 | 152.30 | **0.388** |
| **Fiscal Year** | 2016–2017 | 348 | 0.2227 | 77.50 |  |
|  | 2017–2018 | 1,866 | 0.0829 | 154.71 |  |
|  | 2018–2019 | 2,010 | 0.0519 | 104.36 |  |
|  | 2019–2020 | 1,762 | 0.0527 | 92.85 |  |
|  | 2020–2021 | 1,970 | 0.0626 | 123.25 |  |
|  | 2021–2022 | 1,946 | 0.0785 | 152.80 | **<0.001** |
| **Episode**<br>**persistence** | Continued into ≥next year | 2,271 | 0.1180 | 268.06 |  |
|  | Started+ended same year<br><2022 | 7,631 | 0.0573 | 437.40 | **<0.001** |

Older adults constituted the largest age category, with 2,065 patients aged 60–69 years (20.9%) and 1,863 patients aged ≥ 70 years (18.8%). Children aged 0–9 years comprised 22.4% (n = 2,216) of the total sample. Most cardiovascular medicine users were insured by the NHIF (Table 4).

The mean cardiovascular medicine utilisation (DDD per 100 bed-days) differed across the subgroups. Female patients contributed a larger cumulative DDD load than males (384.61 vs. 320.85 DDD/100 bed-days), and NHIF beneficiaries accounted for the largest cumulative DDD load among the other sponsor categories (446.47 DDD/100 bed-days).

Statistical testing did not demonstrate statistically significant differences across sex or sponsor groups (p > 0.05), whereas the year-to-year variation in DDD per 100 bed-days was significant (p < 0.001).

Across the six fiscal years, the cumulative exposure to cardiovascular medicine totalled 705.46 DDD per 100 bed-days (Figure 2).

**Figure 2:**
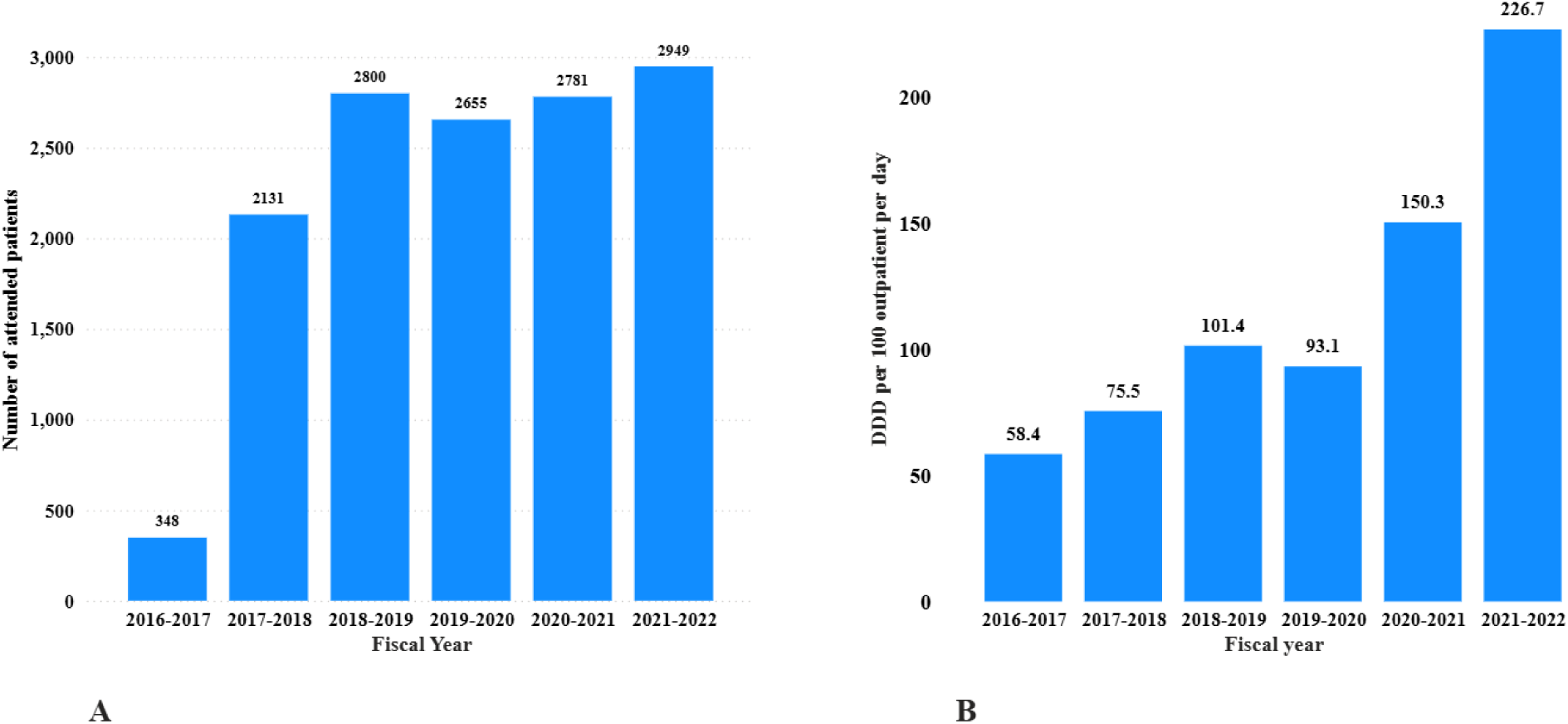
Annual trends in cardiovascular medicine utilisation among inpatients at the Jakaya Kikwete Cardiac Institute (JKCI), Tanzania (2016/2017–2021/2022).

The number of inpatients receiving one or more cardiovascular medicines increased from 348 in 2016/2017 to 2,800 in 2018/2019 and then remained relatively stable until 2021/2022 (N = 2,949) (Figure 2A). Over the same period, the intensity of cardiovascular medicine utilisation also rose, increasing from 58.44 DDD/100 bed-days in 2016/2017 to a peak of 226.74 DDD/100 bed-days in 2021/2022 (Figure 2B). Overall, these longitudinal patterns indicate a sustained upward trajectory of inpatient cardiovascular medicine utilisation over the years.

### Panel A: Annual number of unique inpatients receiving ≥1 cardiovascular medicine (ATC class C). Panel B: Annual utilisation intensity expressed as DDD per 100 bed days

Annual utilisation at ATC Level-3 showed differentiated temporal patterns, with selective calcium channel blockers (C08C) dominating early in the series, while “other cardiac preparations” (C01E) rose sharply toward 2021/2022. Lipid-modifying agents (C10A) and high-ceiling diuretics (C03C) also demonstrated sustained annual growth (Figure 3).

**Figure 3:**
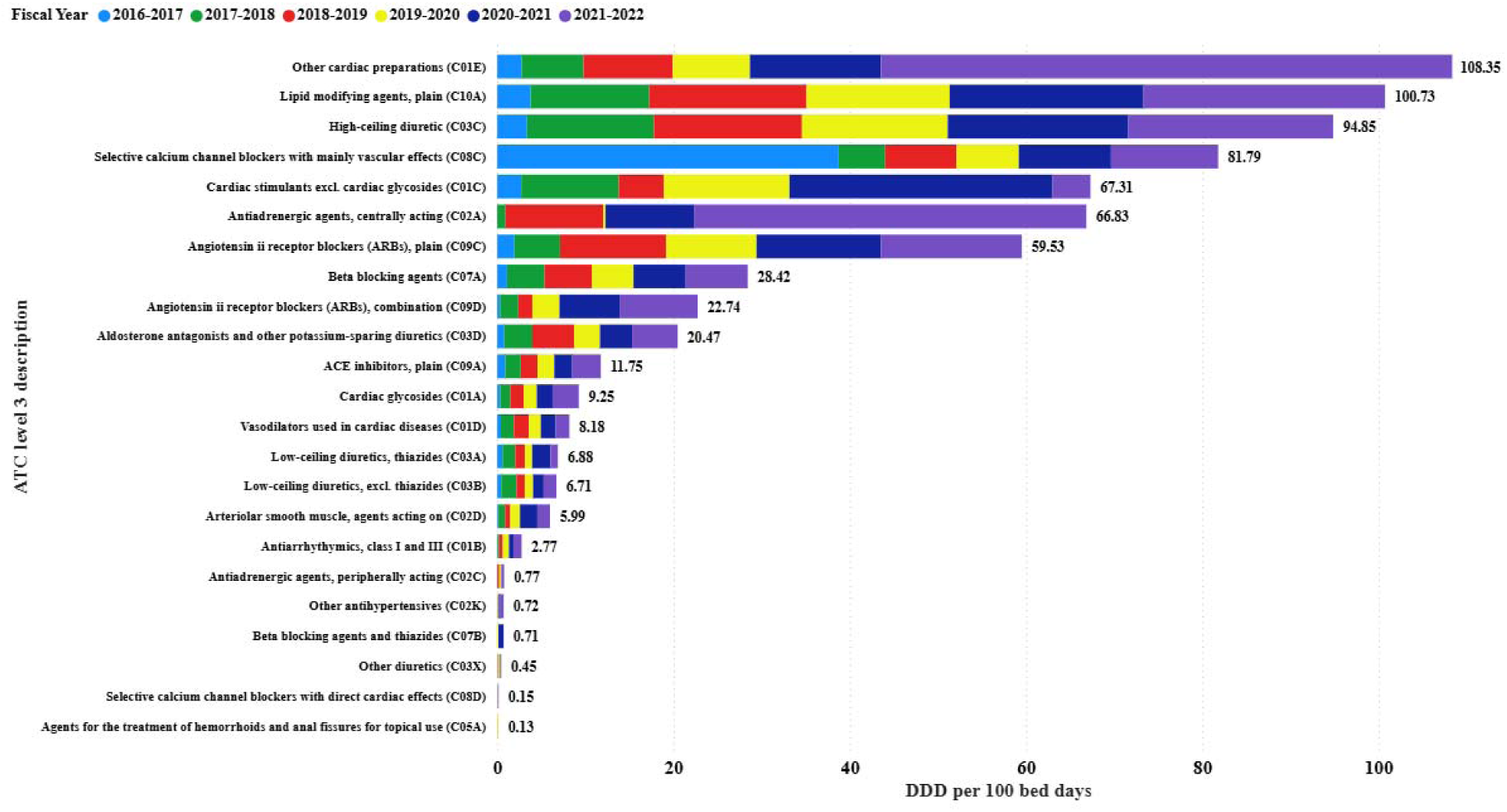
Annual cardiovascular medicine utilisation by ATC level 3 subclasses among inpatients at the Jakaya Kikwete Cardiac Institute (JKCI), Tanzania (2016/2017–2021/2022). Bars represent DDD per 100 bed days per fiscal year, with the rightmost cluster representing cumulative totals across the six-year study period.

Similarly, consistent with the ATC Level 3 patterns, annual utilisation at the molecule level (ATC Level 5) also showed a marked differentiation across agents (Figure 4). Ibuprofen (C01EB16) contributed the highest cumulative DDD/100 bed-days volume within the DU90 segment, whereas norepinephrine (C01CA03) represented the lowest utilisation in this group. Notably, ibuprofen under ATC code C01EB16 corresponds to neonatal ductus-arteriosus closure (cardiac therapeutic indication) rather than conventional analgesic use, and the WHO ATC/DDD index assigns a *course-based* DDD for this molecule.

**Figure 4:**
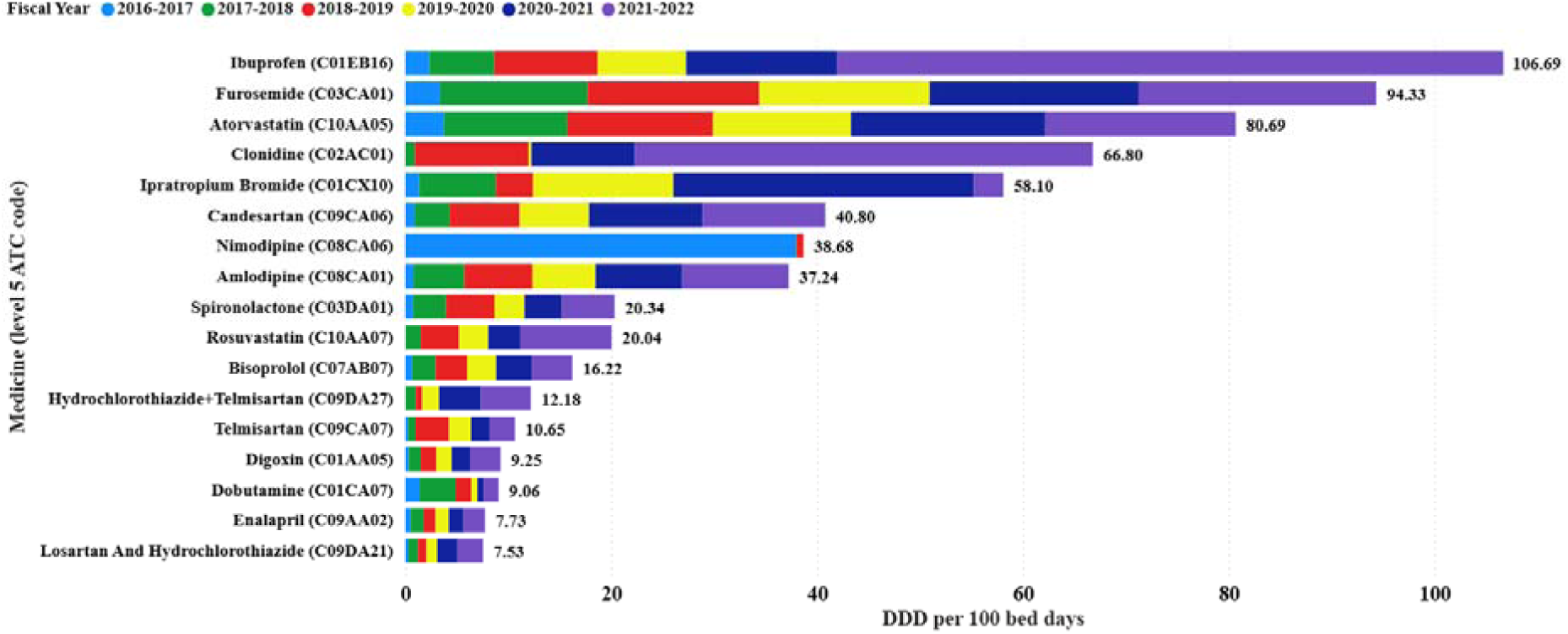
Trends in annual cardiovascular medicine utilisation by ATC Level-5 molecules among inpatients at the Jakaya Kikwete Cardiac Institute (JKCI), Tanzania (2016/2017–2021/2022). The bars represent DDD per 100 bed-days per fiscal year; these 17 molecules constituted the DU90 segment over the six years. Note: Ibuprofen (C01EB16) is coded under cardiac preparations for neonatal ductus arteriosus closure and has a course-based WHO DDD.

Within the ATC Level 5 cardiovascular group, cumulative utilisation also showed a wide variation between individual molecules. Candesartan (C09CA06) was among the least utilised agents, whereas several cardiovascular combinations, particularly telmisartan–hydrochlorothiazide (C09DA27), were among the higher-consumption cardiovascular products. The year-on-year utilisation of these ARB-linked molecules also increased progressively from 2017 to 2021 (Supplementary Table 2).

### Seasonality and longitudinal trend analysis

Monthly cardiovascular medicine utilisation (DDD per 100 bed-days) was further examined to assess seasonal variation and long-term trends. No recurrent seasonality was visually apparent or statistically supported (Kruskal–Wallis p = 0.75), whereas a consistent upward temporal trend was evident (Mann–Kendall Z = 5.82, p < 0.001) (Figure 5; Supplementary Table 3).

**Figure 5.**
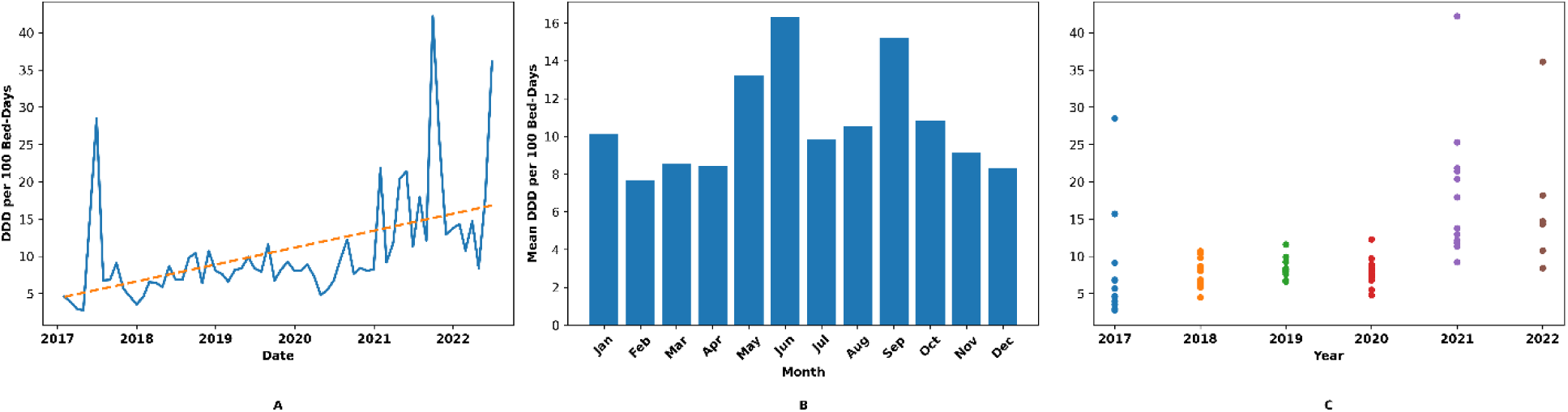
Monthly cardiovascular medicine utilisation among inpatients at the Jakaya Kikwete Cardiac Institute (JKCI), Tanzania (January 2017 – June 2022), expressed as DDD per 100 bed-days.

Panel A: Monthly values with a fitted linear trend line. Panel B: Mean utilisation by calendar month (across years) demonstrating no recurrent seasonal pattern. Panel C: Yearly distribution of monthly DDD values demonstrating increasing utilisation intensity over time.

## Discussion

### Principal findings

This six-year pharmacoepidemiological analysis is the first longitudinal quantification of cardiovascular medicine utilisation in Tanzania using the WHO ATC/DDD methodology. Cardiovascular agents accounted for almost half of all inpatient prescriptions at the Jakaya Kikwete Cardiac Institute (JKCI), with utilisation intensity increasing steadily from 2016 to 2022. Diuretics, particularly furosemide (C03CA01) and spironolactone (C03DA01), formed the dominant therapeutic backbone, reflecting the large heart failure and hypertensive case mix managed at JKCI. Older adults (>60 years) formed the largest user segment, consistent with the known age-related increase in the burden of noncommunicable diseases and clustering of comorbidities.

### Comparison with previous studies

Our findings align with reports from India, Nigeria and other lower-middle-income countries, where high-ceiling and potassium-sparing diuretics remain the first-line therapies for both acute and chronic cardiac care [2,6,16]. The relatively lower consumption of renin–angiotensin system inhibitors and fixed-dose antihypertensive combinations may reflect availability constraints, cost considerations, or prescribing preference. These patterns have been previously reported in similar referral-hospital settings [5,16]. The higher utilisation intensity among older adults is consistent with a prior study linking age, postmenopausal physiology, and metabolic risk clustering with increased VD treatment exposure [8–10]. The present results reinforce the global trends reported by Wylie et al. [1] and Wallace et al. [7], indicating that polypharmacy and chronic medication exposure increase with advancing age.

Comparable prescription profiles dominated by diuretics and lipid-modifying agents in intensive cardiac care have also been described by Nooreen et al. [2], supporting external consistency in a similar critical care cardiology population. Conversely, the relatively limited use of angiotensin receptor blockers and thiazide combinations in the current analysis suggests potential gaps in adherence to the WHO and national treatment guidelines [16].

Notably, ibuprofen coded under C01EB16 was classified under “other cardiac preparations” for neonatal ductus arteriosus closure rather than analgesia; the WHO ATC/DDD index assigns a course-based DDD for this indication.

### Gender, safety and stewardship considerations

Women represented a larger proportion of cardiovascular medicine recipients. This remains biologically coherent: hormonal changes and the transition to peri-and postmenopausal status amplify metabolic risk, endothelial dysfunction and CVD burden [8–10]. Familial predisposition, childhood obesity and metabolic syndrome remain important determinants of lifetime CVD progression [8,11–13]. The existing literature suggests that chronic treatment exposure, including prolonged diuretic use in kidney disease, may further influence cardiovascular risk trajectories [17], although sex-specific pharmacological response differences remain insufficiently characterised [18].

Polypharmacy remains a persistent challenge in tertiary cardiology stewardship. Reducing the number of agents per prescription, reinforcing evidence-based selection, and intensifying monitoring may minimise adverse drug reactions (ADRs), which arise from a complex interplay between medicine-, patient and disease-level drivers [19,20]. This study contributes to the call to increase research in cardiovascular sciences [21] to examine improvements in cardiology medication-prescribing practices.

### Strengths and limitations

This study leveraged a large, continuous, multi-year dataset (>224,000 eligible inpatient prescriptions) and applied standardised WHO ATC/DDD methods, enabling international comparability. In addition, DDD normalisation per 100 bed-days allows signal detection independent of fluctuations in bed occupancy.

However, this was a retrospective, single-centre study, and the causal relationship between prescribing and outcomes could not be assessed. The absence of diagnosis codes, comorbidity variables, and laboratory linkages prevented indication-specific utilisation evaluation. Finally,

JKCI is a national cardiac referral centre, and its utilisation patterns may not accurately reflect those in district or primary care settings.

### Implications for clinicians and policymakers

Routine ATC/DDD reporting could be integrated into hospital stewardship dashboards to support procurement forecasting, formulary decisions, consumption benchmarking and alignment with Tanzania’s Essential Medicines List. This baseline also provides a denominator against which to assess the impact of the National Noncommunicable Disease Strategy and financing reforms through the National Health Insurance Fund.

Although the WHO AWaRe framework currently applies to antibacterials [5], a simple analogous stratification for cardiovascular pharmacotherapy, such as essential/selective/specialised, could support the prioritisation of high-value medicines, reduce unnecessary expenditures, and strengthen accountability in tertiary cardiac care.

### Unanswered questions and future research

Future studies should expand to multicentre cohorts, incorporate diagnosis linkages, and evaluate clinical outcomes (e.g. blood pressure control, readmissions, and mortality). A cost-effectiveness comparison of diuretics, ACE inhibitors, and fixed-dose combinations would strengthen the rationale for procurement. Integrating DDD-based utilisation metrics into electronic medical record systems would enable near-real-time consumption monitoring and feedback to prescribers.

## Conclusion

This study revealed a substantial and increasing intensity of cardiovascular medicine use among inpatients at JKCI from 2016 to 2022, with cardiovascular medicines accounting for nearly half of all prescriptions. Diuretics, statins, and renin-angiotensin system inhibitors have emerged as the most frequently prescribed agents, underscoring their central role in the pharmacological management of cardiovascular conditions.

These prescribing patterns have important implications for the future of cardiovascular care in Tanzania. The growing demand highlights the urgency of strengthening stewardship programs, procurement mechanisms, and supply chain systems to ensure reliable access to essential medicines. Although the predominance of guideline-recommended therapies reflects broadly appropriate prescribing, it also highlights the need for regular prescription audits, focused prescriber training, and the adoption of digital prescribing tools to promote rational prescribing.

The concentration of prescriptions among a relatively small group of medicines further highlights the need for ongoing monitoring of adverse events, treatment effectiveness, and economic impact. These findings offer a data-driven foundation for national policy, clinical governance, and quality improvement in cardiovascular care, particularly in resource-constrained settings.

Sustained pharmacoepidemiological surveillance is essential to guide evidence-based interventions and enhance the quality, safety, and efficiency of cardiovascular medicines used in Tanzania’s health system.

## Data Availability Statement

Due to patient confidentiality restrictions and the Jakaya Kikwete Cardiac Institute’s (JKCI) institutional ownership of the data, the dataset cannot be shared publicly. Aggregated de-identified datasets may be provided by the corresponding author upon reasonable request and with permission from the JKCI.

## Ethics approval

This retrospective analysis of de-identified inpatient dispensing data was approved by the Muhimbili University of Health and Allied Sciences Research Ethics Committee (Ref: DA.25/111/01B/184). The requirement for informed consent was waived because the dataset contained no direct personal identifiers. Data processing steps (Power Query transformation, ATC-DDD mapping, and removal of personal identifiers) were performed in a secure JKCI environment prior to export.

## Author Contributions

- **Philip Sasi:** Data interpretation, literature review and writing the first draft.
- **Justine L. Vasco:** Data extraction, cleaning, statistical analysis, drafting of the manuscript.
- **Raphael Z. Sangeda:** Conceptualisation, study design, supervision, data validation, critical review, and corresponding author responsibilities.
- **Tusaligwe Mbilinyi:** Data cleaning and statistical analysis
- **Naizihijwa Majani:** Data access, quality control, and institutional liaison at JKCI.
- **Peter Richard Kisenge:** Clinical oversight, manuscript review and policy context integration.
- **Pilly Chillo:** Clinical oversight, manuscript review
- All authors have approved the final version of the manuscript.

## Acknowledgements

We acknowledge the management and data staff of the Jakaya Kikwete Cardiac Institute for facilitating access to and cleaning data.

