## Supplementary Tables for "Pharmacoepidemiology of Cardiovascular Medicines Used Among Inpatients at a Tertiary Care Hospital in Tanzania, 2016-2022"

**Supplementary Materials**

**Supplementary Table 1: Composition of prescription records for all medicines-utilisation-distribution at ATC level 5**

| **Rank** | **Generic name ( ATC level 5 code)** | **Number of records** | **Cumulative records** | **Cumulative %** |
| --- | --- | --- | --- | --- |
| 1 | FUROSEMIDE (C03CA01) | 28,861 | 28,861 | 12.9 |
| 2 | SPIRONOLACTONE (C03DA01) | 14,039 | 42,900 | 19.1 |
| 3 | PARACETAMOL (N02BE01) | 13,835 | 56,735 | 25.3 |
| 4 | CEFTRIAXONE (J01DD04) | 8,491 | 65,226 | 29.1 |
| 5 | HEPARIN (B01AB01) | 8,076 | 73,302 | 32.7 |
| 6 | ACETYLSALICYLIC ACID (A01AD05) | 7,118 | 80,420 | 35.9 |
| 7 | PANTOPRAZOLE (A02BC02) | 6,489 | 86,909 | 38.7 |
| 8 | ATORVASTATIN (C10AA05) | 6,453 | 93,362 | 41.6 |
| 9 | ISOSORBIDE DINITRATE (C01DA08) | 5,054 | 98,416 | 43.9 |
| 10 | SODIUM CHLORIDE (A12CA01) | 4,624 | 103,040 | 45.9 |
| 11 | CLOPIDOGREL (B01AC04) | 4,598 | 107,638 | 48.0 |
| 12 | ENALAPRIL (C09AA02) | 4,269 | 111,907 | 49.9 |
| 13 | CARVEDILOL (C07AG02) | 4,208 | 116,115 | 51.8 |
| 14 | BISOPROLOL (C07AB07) | 4,148 | 120,263 | 53.6 |
| 15 | CANDESARTAN (C09CA06) | 3,753 | 124,016 | 55.3 |
| 16 | WARFARIN (B01AA03) | 3,699 | 127,715 | 56.9 |
| 17 | DIGOXIN (C01AA05) | 3,626 | 131,341 | 58.6 |
| 18 | AMLODIPINE (C08CA01) | 2,970 | 134,311 | 59.9 |
| 19 | IBUPROFEN (C01EB16) | 2,922 | 137,233 | 61.2 |
| 20 | AMOXICILLIN + CLAVULANIC (J01CR02) | 2,920 | 140,153 | 62.5 |
| 21 | AMOXICILLIN (J01CA04) | 2,824 | 142,977 | 63.7 |
| 22 | MEROPENEM (J01DH02) | 2,756 | 145,733 | 65.0 |
| 23 | POTASSIUM CHLORIDE (A12BA01) | 2,656 | 148,389 | 66.2 |
| 24 | METRONIDAZOLE (J01XD01) | 2,629 | 151,018 | 67.3 |
| 25 | PIPERACILLIN+TAZOBACTAM (J01CR05) | 2,444 | 153,462 | 68.4 |
| 26 | METOLAZONE (C03BA08) | 2,265 | 155,727 | 69.4 |
| 27 | VANCOMYCIN (J01XA01) | 2,228 | 157,955 | 70.4 |
| 28 | CAPTOPRIL (C09AA01) | 2,123 | 160,078 | 71.4 |
| 29 | METOPROLOL (C07AB02) | 1,869 | 161,947 | 72.2 |
| 30 | MULTIVITAMINS+OTHER COMBINATIONS (A11AB50) | 1,776 | 163,723 | 73.0 |
| 31 | BUDESONIDE (R01AD05) | 1,641 | 165,364 | 73.7 |
| 32 | FERROUS SULPHATE (B03AD03) | 1,599 | 166,963 | 74.4 |
| 33 | ADRENALINE (A01AD06) | 1,576 | 168,539 | 75.1 |
| 34 | ESOMEPRAZOLE (A02BC05) | 1,545 | 170,084 | 75.8 |
| 35 | SALBUTAMOL (R03AC02) | 1,527 | 171,611 | 76.5 |
| 36 | MONTELUKAST (R03DC03) | 1,458 | 173,069 | 77.2 |
| 37 | HYDRALAZINE (C02DB02) | 1,416 | 174,485 | 77.8 |
| 38 | CALCIUM GLUCONATE (A12AA03) | 1,411 | 175,896 | 78.4 |
| 39 | METFORMIN (A10BA02) | 1,293 | 177,189 | 79.0 |
| 40 | AMIODARONE (C01BD01) | 1,286 | 178,475 | 79.6 |
| 41 | PREDNISOLONE (A01AC04) | 1,264 | 179,739 | 80.1 |
| 42 | GLIMEPIRIDE (A10BB12) | 1,225 | 180,964 | 80.7 |
| 43 | MELOXICAM (M01AC06) | 1,182 | 182,146 | 81.2 |
| 44 | OMEPRAZOLE (A02BC01) | 1,157 | 183,303 | 81.7 |
| 45 | GENTAMICIN (J01GB03) | 1,143 | 184,446 | 82.2 |
| 46 | AMPICLOXACILLIN (J01CA51) | 1,052 | 185,498 | 82.7 |
| 47 | DOBUTAMINE (C01CA07) | 982 | 186,480 | 83.1 |
| 48 | SODIUM BICARBONATE (B05CB04) | 973 | 187,453 | 83.6 |
| 49 | RABEPRAZOLE (A02BC04) | 884 | 188,337 | 84.0 |
| 50 | FERROUS FUMARATE (B03AA02) | 879 | 189,216 | 84.4 |
| 51 | VITAMIN (V04CB01) | 829 | 190,045 | 84.7 |
| 52 | PROPRANOLOL (C07AA05) | 792 | 190,837 | 85.1 |
| 53 | ROSUVASTATIN (C10AA07) | 779 | 191,616 | 85.4 |
| 54 | MULTIVITAMINS AND CALCIUM (A11AA02) | 768 | 192,384 | 85.8 |
| 55 | TELMISARTAN (C09CA07) | 756 | 193,140 | 86.1 |
| 56 | PHENOXYMETHYLPENICILLIN-BENZATHINE (J01CE10) | 736 | 193,876 | 86.4 |
| 57 | LOSARTAN AND HYDROCHLOROTHIAZIDE (C09DA21) | 725 | 194,601 | 86.8 |
| 58 | CIPROFLOXACIN (J01MA02) | 725 | 195,326 | 87.1 |
| 59 | LOSARTAN (C09CA01) | 715 | 196,041 | 87.4 |
| 60 | HYDROCHLOROTHIAZIDE+TELMISARTAN (C09DA27) | 686 | 196,727 | 87.7 |
| 61 | AMPICILLIN (J01CA01) | 685 | 197,412 | 88.0 |
| 62 | INSULIN ASPART (A10AB05) | 682 | 198,094 | 88.3 |
| 63 | PREGABALIN (N03AX16) | 658 | 198,752 | 88.6 |
| 64 | AZITHROMYCIN (J01FA10) | 636 | 199,388 | 88.9 |
| 65 | ISOSORBIDE MONONITRATE (C01DA14) | 624 | 200,012 | 89.2 |
| 66 | FOLIC ACID (B03BB01) | 622 | 200,634 | 89.5 |
| 67 | ALUMINIUM HYDROXIDE+MAGNESIUM HYDROXIDE (A02AD10) | 620 | 201,254 | 89.7 |
| 68 | DEXAMETHASONE (A01AC02) | 611 | 201,865 | 90.0 |
| 69 | FLUCONAZOLE (J02AC01) | 591 | 202,456 | 90.3 |
| 70 | RIVAROXABAN (B01AF01) | 582 | 203,038 | 90.5 |
| 71 | TOLVAPTAN (C03XA01) | 559 | 203,597 | 90.8 |
| 72 | SILDENAFIL (C02KX06) | 559 | 204,156 | 91.0 |
| 73 | TRAMADOL (N02AX02) | 557 | 204,713 | 91.3 |
| 74 | FEBUXOSTAT (M04AA03) | 542 | 205,255 | 91.5 |
| 75 | GLIBENCLAMIDE (A10BB01) | 536 | 205,791 | 91.8 |
| 76 | LORATADINE (R06AX13) | 517 | 206,308 | 92.0 |
| 77 | GUAIFENESIN (R05CA03) | 516 | 206,824 | 92.2 |
| 78 | HYDROCORTISONE (A01AC03) | 502 | 207,326 | 92.4 |
| 79 | RANITIDINE (A02BA02) | 501 | 207,827 | 92.7 |
| 80 | CETIRIZIN (R06AE07) | 500 | 208,327 | 92.9 |
| 81 | ALBENDAZOLE (P02CA03) | 460 | 208,787 | 93.1 |
| 82 | LACTULOSE (A06AD11) | 447 | 209,234 | 93.3 |
| 83 | NIFEDIPINE (C08CA05) | 442 | 209,676 | 93.5 |
| 84 | ZINC SULFATE (A12CB01) | 432 | 210,108 | 93.7 |
| 85 | IPRATROPIUM BROMIDE (C01CX10) | 428 | 210,536 | 93.9 |
| 86 | DEXTROSE (B05AA05) | 415 | 210,951 | 94.1 |
| 87 | GLYCERYL TRINITRATE (A05AX08) | 384 | 211,335 | 94.2 |
| 88 | SODIUM LACTATE (B05XA08) | 356 | 211,691 | 94.4 |
| 89 | CEFPODOXIME (J01DD13) | 327 | 212,018 | 94.5 |
| 90 | DESLORATADINE (R06AX27) | 315 | 212,333 | 94.7 |
| 91 | PHENOBARBITAL (N03AA02) | 307 | 212,640 | 94.8 |
| 92 | FLUCLOXACILLIN (J01CF05) | 303 | 212,943 | 94.9 |
| 93 | BENDROFLUMETHIAZIDE (C03AA01) | 296 | 213,239 | 95.1 |
| 94 | METOCLOPRAMIDE (A03FA01) | 291 | 213,530 | 95.2 |
| 95 | MENTHOL (R01AX23) | 287 | 213,817 | 95.3 |
| 96 | IPRATROPIUM BROMIDE+SALBUTAMOL (R03AL02) | 287 | 214,104 | 95.5 |
| 97 | ATROPINE (A03BA01) | 265 | 214,369 | 95.6 |
| 98 | CARBAMAZEPINE (N03AF01) | 264 | 214,633 | 95.7 |
| 99 | MAGNESIUM SULFATE (A06AD04) | 261 | 214,894 | 95.8 |
| 100 | HALOPERIDOL (N05AD01) | 260 | 215,154 | 95.9 |
| 101 | ARTESUNATE (P01BE03) | 253 | 215,407 | 96.0 |
| 102 | LEVOTHYROXINE (H03AA01) | 249 | 215,656 | 96.2 |
| 103 | AMITRIPTYLINE (N06AA09) | 230 | 215,886 | 96.3 |
| 104 | TAMSULOSIN (G04CA02) | 228 | 216,114 | 96.4 |
| 105 | MIDAZOLAM (N03AE02) | 228 | 216,342 | 96.5 |
| 106 | ARTEMETHER (P01BE02) | 226 | 216,568 | 96.6 |
| 107 | CEFIXIME (J01DD08) | 218 | 216,786 | 96.7 |
| 108 | ASPRIN+CLOPIDOGREL (B01AC34) | 216 | 217,002 | 96.8 |
| 109 | CLARITHROMYCIN (J01FA09) | 209 | 217,211 | 96.8 |
| 110 | CEFALEXIN (J01DB01) | 206 | 217,417 | 96.9 |
| 111 | IRBESARTAN (C09CA04) | 205 | 217,622 | 97.0 |
| 112 | ALLOPURINOL (M04AA01) | 192 | 217,814 | 97.1 |
| 113 | ARTEMETHER + LUMEFANTRINE (P01BF01) | 191 | 218,005 | 97.2 |
| 114 | DOXAZOSIN (C02CA04) | 189 | 218,194 | 97.3 |
| 115 | NEBIVOLOL (C07AB12) | 187 | 218,381 | 97.4 |
| 116 | VILDAGLIPTIN (A10BH02) | 181 | 218,562 | 97.4 |
| 117 | BETAHISTINE (N07CA01) | 179 | 218,741 | 97.5 |
| 118 | GLIBENCLAMID+METFORMIN (A10BD31) | 172 | 218,913 | 97.6 |
| 119 | DICLOFENAC (M01AB05) | 169 | 219,082 | 97.7 |
| 120 | ACETAZOLAMIDE (S01EC01) | 163 | 219,245 | 97.8 |
| 121 | BISACODYL (A06AB02) | 153 | 219,398 | 97.8 |
| 122 | DIAZEPAM (N05BA01) | 145 | 219,543 | 97.9 |
| 123 | FINASTERIDE (G04CB01) | 137 | 219,680 | 97.9 |
| 124 | ALBUMIN (B05AA01) | 136 | 219,816 | 98.0 |
| 125 | PHYTOMENADIONE (B02BA01) | 135 | 219,951 | 98.1 |
| 126 | MEBENDAZOLE (P02CA01) | 132 | 220,083 | 98.1 |
| 127 | FELODIPINE (C08CA02) | 116 | 220,199 | 98.2 |
| 128 | LISINOPRIL (C09AA03) | 112 | 220,311 | 98.2 |
| 129 | DOMPERIDONE (A03FA03) | 111 | 220,422 | 98.3 |
| 130 | THIAMINE (N07XB52) | 108 | 220,530 | 98.3 |
| 131 | VERAPAMIL (C08DA01) | 107 | 220,637 | 98.4 |
| 132 | TRANEXAMIC ACID (B02AA02) | 107 | 220,744 | 98.4 |
| 133 | ONDANSETRON (A04AA01) | 107 | 220,851 | 98.5 |
| 134 | MULTIVITAMINS AND OTHER MINERALS, INCL. COMBINATIONS (A11AA03) | 107 | 220,958 | 98.5 |
| 135 | RANOLAZINE (C01EB18) | 105 | 221,063 | 98.6 |
| 136 | GABAPENTIN (N03AX12) | 102 | 221,165 | 98.6 |
| 137 | CANDESARTAN+HYDROCHLOROTHIAZIDE (C09DA26) | 101 | 221,266 | 98.7 |
| 138 | SACCHARATED IRON OXIDE (B03AB02) | 96 | 221,362 | 98.7 |
| 139 | ACICLOVIR (J05AB01) | 96 | 221,458 | 98.7 |
| 140 | POVIDONE IODINE (S01AX18) | 94 | 221,552 | 98.8 |
| 141 | AMLODIPINE+HYDROCHLOROTHIAZIDE+VALSARTAN (C09DX01) | 92 | 221,644 | 98.8 |
| 142 | DOPAMINE (C01CA04) | 91 | 221,735 | 98.9 |
| 143 | DAPAGLIFLOZIN (A10BK01) | 83 | 221,818 | 98.9 |
| 144 | TORSEMIDE (C03CA04) | 79 | 221,897 | 98.9 |
| 145 | IVABRADINE (C01EB17) | 78 | 221,975 | 99.0 |
| 146 | SITAGLIPTIN (A10BH01) | 77 | 222,052 | 99.0 |
| 147 | VALSARTAN AND SACUBITRIL (C09DX04) | 75 | 222,127 | 99.0 |
| 148 | FLUTICASONE+SALMETEROL (R03AK06) | 75 | 222,202 | 99.1 |
| 149 | SULFAMETHOXAZOLE+TRIMETHOPRIM (J01EE01) | 74 | 222,276 | 99.1 |
| 150 | DOXYCYCLINE (J01AA02) | 74 | 222,350 | 99.1 |
| 151 | ADENOSINE (C01EB10) | 71 | 222,421 | 99.2 |
| 152 | KETOCONAZOLE (D01AC08) | 65 | 222,486 | 99.2 |
| 153 | LIDOCAINE (A01AE01) | 64 | 222,550 | 99.2 |
| 154 | TERBINAFINE (D01AE15) | 63 | 222,613 | 99.3 |
| 155 | ATENOLOL (C07AB03) | 63 | 222,676 | 99.3 |
| 156 | TRIMETAZIDINE (C01EB15) | 60 | 222,736 | 99.3 |
| 157 | LOPERAMIDE (A07DA03) | 60 | 222,796 | 99.3 |
| 158 | CARBIMAZOLE (H03BB01) | 58 | 222,854 | 99.4 |
| 159 | EPLERENONE (C03DA04) | 57 | 222,911 | 99.4 |
| 160 | STREPTOKINASE (B01AD01) | 52 | 222,963 | 99.4 |
| 161 | BACLOFEN (M03BX01) | 51 | 223,014 | 99.4 |
| 162 | MILRINONE (C01CE02) | 50 | 223,064 | 99.5 |
| 163 | BOSENTAN (C02KX01) | 49 | 223,113 | 99.5 |
| 164 | FERROUS SULPHATE+FOLIC ACID (B03BB51) | 48 | 223,161 | 99.5 |
| 165 | TINIDAZOLE (J01XD02) | 45 | 223,206 | 99.5 |
| 166 | LEVODOPA (N04BA01) | 45 | 223,251 | 99.5 |
| 167 | CLONIDINE (C02AC01) | 44 | 223,295 | 99.6 |
| 168 | PIOGLITAZONE (A10BG03) | 41 | 223,336 | 99.6 |
| 169 | MANNITOL (A06AD16) | 41 | 223,377 | 99.6 |
| 170 | CEFUROXIME (J01DC02) | 41 | 223,418 | 99.6 |
| 171 | FLUTICASONE (D07AC17) | 39 | 223,457 | 99.6 |
| 172 | AMIKACIN (J01GB06) | 36 | 223,493 | 99.6 |
| 173 | INSULIN (HUMAN) (A10AB01) | 35 | 223,528 | 99.7 |
| 174 | GLUCOSAMINE (M01AX05) | 33 | 223,561 | 99.7 |
| 175 | LORAZEPAM (N05BA06) | 32 | 223,593 | 99.7 |
| 176 | TRIHEXYPHENIDYL (N04AA01) | 31 | 223,624 | 99.7 |
| 177 | ATRACURIUM (M03AC04) | 29 | 223,653 | 99.7 |
| 178 | BROMAZEPAM (N05BA08) | 27 | 223,680 | 99.7 |
| 179 | LIGNOCAINE (N01BB02) | 24 | 223,704 | 99.7 |
| 180 | CEFTRIAXONE AND BETA-LACTAMASE INHIBITOR (J01DD63) | 24 | 223,728 | 99.7 |
| 181 | CEFOPERAZONE+SALBACTAM (J01DD62) | 24 | 223,752 | 99.8 |
| 182 | ORNIDAZOLE (G01AF06) | 21 | 223,773 | 99.8 |
| 183 | NOREPINEPHRINE (C01CA03) | 21 | 223,794 | 99.8 |
| 184 | ARTEMISININ PIPERAQUINE (P01BF07) | 21 | 223,815 | 99.8 |
| 185 | CHLORPROMAZINE (N05AA07) | 20 | 223,835 | 99.8 |
| 186 | HYOSCINE BUTYLBROMIDE (A03BB01) | 19 | 223,854 | 99.8 |
| 187 | DILTIAZEM (C05AE03) | 19 | 223,873 | 99.8 |
| 188 | AMINOPHYLLINE (R03DA05) | 18 | 223,891 | 99.8 |
| 189 | GLYCEROL (A06AG04) | 17 | 223,908 | 99.8 |
| 190 | PHENYTOIN (N03AB02) | 16 | 223,924 | 99.8 |
| 191 | KETAMINE (N01AX03) | 16 | 223,940 | 99.8 |
| 192 | OCTREOTIDE (H01CB02) | 14 | 223,954 | 99.9 |
| 193 | NYSTATIN (A01AB33) | 14 | 223,968 | 99.9 |
| 194 | MICONAZOLE (J02AB01) | 14 | 223,982 | 99.9 |
| 195 | DOXORUBICIN (L01DB01) | 14 | 223,996 | 99.9 |
| 196 | COLCHICINE (M04AC01) | 14 | 224,010 | 99.9 |
| 197 | ASCORBIC ACID (G01AD03) | 14 | 224,024 | 99.9 |
| 198 | ZOFENOPRIL (C09AA15) | 12 | 224,036 | 99.9 |
| 199 | TENECTEPLASE (B01AD11) | 12 | 224,048 | 99.9 |
| 200 | KETOPROFEN (M01AE03) | 12 | 224,060 | 99.9 |
| 201 | CEFEPIME (J01DE01) | 11 | 224,071 | 99.9 |
| 202 | ARTEMISININ (P01BE01) | 11 | 224,082 | 99.9 |
| 203 | IVERMECTIN (P02CF01) | 10 | 224,092 | 99.9 |
| 204 | RAMIPRIL (C09AA05) | 9 | 224,101 | 99.9 |
| 205 | NORETHISTERONE (G03AC01) | 9 | 224,110 | 99.9 |
| 206 | NICORANDIL (C01DX16) | 9 | 224,119 | 99.9 |
| 207 | ERYTHROMYCIN (J01FA01) | 9 | 224,128 | 99.9 |
| 208 | AMOXICILLIN+METRONIDAZOLE+OMEPRAZOLE (A02BD01) | 9 | 224,137 | 99.9 |
| 209 | PROPOFOL (N01AX10) | 8 | 224,145 | 99.9 |
| 210 | PHENOXYMETHYLPENICILLIN (J01CE02) | 8 | 224,153 | 99.9 |
| 211 | MORPHINE (N02AA01) | 8 | 224,161 | 99.9 |
| 212 | BROMOCRIPTINE (N04BC01) | 8 | 224,169 | 99.9 |
| 213 | ACETYLCYSTEININE (V03AB23) | 7 | 224,176 | 99.9 |
| 214 | METHYLPREDNISOLONE (D07AA01) | 6 | 224,182 | 100.0 |
| 215 | METHYLDOPA (LEVOROTATORY) (C02AB01) | 6 | 224,188 | 100.0 |
| 216 | ITRACONAZOLE (J02AC02) | 6 | 224,194 | 100.0 |
| 217 | GLICLAZIDE (A10BB09) | 6 | 224,200 | 100.0 |
| 218 | DIPHENHYDRAMINE (A04AB05) | 6 | 224,206 | 100.0 |
| 219 | ACECLOFENAC (M01AB16) | 6 | 224,212 | 100.0 |
| 220 | NEBIVOLOL+THIAZIDE (C07BB12) | 5 | 224,217 | 100.0 |
| 221 | CALCIUM CARBONATE (A02AC01) | 5 | 224,222 | 100.0 |
| 222 | TOCOPHEROL (VITAMIN E) (A11HA03) | 4 | 224,226 | 100.0 |
| 223 | NIMODIPINE (C08CA06) | 4 | 224,230 | 100.0 |
| 224 | LEVETIRACETAM (N03AX14) | 4 | 224,234 | 100.0 |
| 225 | VALSARTAN (C09CA03) | 3 | 224,237 | 100.0 |
| 226 | PHENYLEPHRINE (C01CA06) | 3 | 224,240 | 100.0 |
| 227 | PANCURONIUM (M03AC01) | 3 | 224,243 | 100.0 |
| 228 | METHYLEPREDINISOLONE (H02AB04) | 3 | 224,246 | 100.0 |
| 229 | LABETALOL (C07AG01) | 3 | 224,249 | 100.0 |
| 230 | IPRATROPIUM (R01AX03) | 3 | 224,252 | 100.0 |
| 231 | FLUOXETINE (N06AB03) | 3 | 224,255 | 100.0 |
| 232 | CLINDAMYCIN (J01FF01) | 3 | 224,258 | 100.0 |
| 233 | TRIAMCINOLONE (A01AC01) | 2 | 224,260 | 100.0 |
| 234 | SODIUM CITRATE (B05CB02) | 2 | 224,262 | 100.0 |
| 235 | REMDESIVIR (J05AB16) | 2 | 224,264 | 100.0 |
| 236 | PYRIMETHAMINE (P01BD01) | 2 | 224,266 | 100.0 |
| 237 | IMIPENEM+CILASTATIN (J01DH51) | 2 | 224,268 | 100.0 |
| 238 | FORMOTEROL AND BUDESONIDE (R03AK07) | 2 | 224,270 | 100.0 |
| 239 | DICLOFENAC+PARACETAMOL+SERRATIOPEPTIDASE (M01AB55) | 2 | 224,272 | 100.0 |
| 240 | CEFADROXIL (J01DB05) | 2 | 224,274 | 100.0 |
| 241 | BECLOMETHASONE + SALBUTAMOL (R03BA01) | 2 | 224,276 | 100.0 |
| 242 | PROTAMINE (V03AB14) | 1 | 224,277 | 100.0 |
| 243 | PROMETHAZINE (N05CM22) | 1 | 224,278 | 100.0 |
| 244 | PARACETAMOL, COMBINATION (N02CX63) | 1 | 224,279 | 100.0 |
| 245 | OLMESARTAN MEDOXOMIL (C09CA08) | 1 | 224,280 | 100.0 |
| 246 | NICOTINE (N07BA01) | 1 | 224,281 | 100.0 |
| 247 | NEOSTIGMINE (N07AA01) | 1 | 224,282 | 100.0 |
| 248 | LIQUID PARAFFIN (A06AA01) | 1 | 224,283 | 100.0 |
| 249 | INDAPAMIDE (C03BA11) | 1 | 224,284 | 100.0 |
| 250 | HYDROCODONE+PARACETAMOL (N02AJ01) | 1 | 224,285 | 100.0 |
| 251 | GRISEOFULVIN (D01AA08) | 1 | 224,286 | 100.0 |
| 252 | FLECAINIDE (C01BC04) | 1 | 224,287 | 100.0 |
| 253 | EPHEDRINE (C01CA26) | 1 | 224,288 | 100.0 |
| 254 | EMPAGLIFLOZIN (A10BK03) | 1 | 224,289 | 100.0 |
| 255 | CINNARIZINE (N06DX12) | 1 | 224,290 | 100.0 |
|  | **Total** | **224,290** |  |  |

**Supplementary Table 2: Composition of prescription records for cardiovascular (class C) medicines-utilisation-distribution at ATC level 5**

| **Medicine (ATC level 5 code)** | **DDD per 100 bed-days** | **Percent** | **Cumulative Percent** | **Rank** | **DU zone** |
| --- | --- | --- | --- | --- | --- |
| Ibuprofen (C01EB16) | 106.6864 | 15.1 | 15.1 | 1 | DU50 |
| Furosemide (C03CA01) | 94.3288 | 13.4 | 28.5 | 2 | DU50 |
| Atorvastatin (C10AA05) | 80.6871 | 11.4 | 39.9 | 3 | DU50 |
| Clonidine (C02AC01) | 66.802 | 9.5 | 49.4 | 4 | DU50 |
| Ipratropium Bromide (C01CX10) | 58.1025 | 8.2 | 57.6 | 5 | DU90 |
| Candesartan (C09CA06) | 40.7959 | 5.8 | 63.4 | 6 | DU90 |
| Nimodipine (C08CA06) | 38.6835 | 5.5 | 68.9 | 7 | DU90 |
| Amlodipine (C08CA01) | 37.2392 | 5.3 | 74.2 | 8 | DU90 |
| Spironolactone (C03DA01) | 20.336 | 2.9 | 77.1 | 9 | DU90 |
| Rosuvastatin (C10AA07) | 20.041 | 2.8 | 79.9 | 10 | DU90 |
| Bisoprolol (C07AB07) | 16.2175 | 2.3 | 82.2 | 11 | DU90 |
| Hydrochlorothiazide+Telmisartan (C09DA27) | 12.1813 | 1.7 | 83.9 | 12 | DU90 |
| Telmisartan (C09CA07) | 10.6476 | 1.5 | 85.4 | 13 | DU90 |
| Digoxin (C01AA05) | 9.2518 | 1.3 | 86.8 | 14 | DU90 |
| Dobutamine (C01CA07) | 9.0553 | 1.3 | 88.0 | 15 | DU90 |
| Enalapril (C09AA02) | 7.7254 | 1.1 | 89.1 | 16 | DU90 |
| Losartan And Hydrochlorothiazide (C09DA21) | 7.533 | 1.1 | 90.2 | 17 | DU90 |
| Bendroflumethiazide (C03AA01) | 6.8778 | 1.0 | 91.2 | 18 |  |
| Isosorbide Dinitrate (C01DA08) | 6.7671 | 1.0 | 92.1 | 19 |  |
| Metolazone (C03BA08) | 6.7085 | 1.0 | 93.1 | 20 |  |
| Carvedilol (C07AG02) | 6.2768 | 0.9 | 94.0 | 21 |  |
| Hydralazine (C02DB02) | 5.9899 | 0.8 | 94.8 | 22 |  |
| Losartan (C09CA01) | 4.7638 | 0.7 | 95.5 | 23 |  |
| Nifedipine (C08CA05) | 3.883 | 0.6 | 96.0 | 24 |  |
| Metoprolol (C07AB02) | 3.1947 | 0.5 | 96.5 | 25 |  |
| Irbesartan (C09CA04) | 3.1852 | 0.5 | 97.0 | 26 |  |
| Captopril (C09AA01) | 3.1161 | 0.4 | 97.4 | 27 |  |
| Amiodarone (C01BD01) | 2.7658 | 0.4 | 97.8 | 28 |  |
| Felodipine (C08CA02) | 1.9853 | 0.3 | 98.1 | 29 |  |
| Nebivolol (C07AB12) | 1.6647 | 0.2 | 98.3 | 30 |  |
| Isosorbide Mononitrate (C01DA14) | 1.4116 | 0.2 | 98.5 | 31 |  |
| Candesartan+Hydrochlorothiazide (C09DA26) | 1.3108 | 0.2 | 98.7 | 32 |  |
| Amlodipine+Hydrochlorothiazide+Valsartan (C09DX01) | 1.2758 | 0.2 | 98.9 | 33 |  |
| Trimetazidine (C01EB15) | 1.1065 | 0.2 | 99.0 | 34 |  |
| Propranolol (C07AA05) | 0.8496 | 0.1 | 99.1 | 35 |  |
| Doxazosin (C02CA04) | 0.7701 | 0.1 | 99.3 | 36 |  |
| Nebivolol+Thiazide (C07BB12) | 0.7096 | 0.1 | 99.4 | 37 |  |
| Sildenafil (C02KX06) | 0.5613 | 0.1 | 99.4 | 38 |  |
| Torsemide (C03CA04) | 0.5196 | 0.1 | 99.5 | 39 |  |
| Tolvaptan (C03XA01) | 0.4463 | 0.1 | 99.6 | 40 |  |
| Valsartan And Sacubitril (C09DX04) | 0.4344 | 0.1 | 99.6 | 41 |  |
| Lisinopril (C09AA03) | 0.4142 | 0.1 | 99.7 | 42 |  |
| Ranolazine (C01EB18) | 0.4062 | 0.1 | 99.8 | 43 |  |
| Ramipril (C09AA05) | 0.4059 | 0.1 | 99.8 | 44 |  |
| Atenolol (C07AB03) | 0.2197 | 0.0 | 99.8 | 45 |  |
| Bosentan (C02KX01) | 0.155 | 0.0 | 99.9 | 46 |  |
| Dopamine (C01CA04) | 0.1516 | 0.0 | 99.9 | 47 |  |
| Verapamil (C08DA01) | 0.1497 | 0.0 | 99.9 | 48 |  |
| Eplerenone (C03DA04) | 0.1297 | 0.0 | 99.9 | 49 |  |
| Diltiazem (C05AE03) | 0.1292 | 0.0 | 99.9 | 50 |  |
| Ivabradine (C01EB17) | 0.118 | 0.0 | 100.0 | 51 |  |
| Valsartan (C09CA03) | 0.1084 | 0.0 | 100.0 | 52 |  |
| Zofenopril (C09AA15) | 0.0849 | 0.0 | 100.0 | 53 |  |
| Methyldopa (Levorotatory) (C02AB01) | 0.0318 | 0.0 | 100.0 | 54 |  |
| Adenosine (C01EB10) | 0.0317 | 0.0 | 100.0 | 55 |  |
| Olmesartan Medoxomil (C09CA08) | 0.0256 | 0.0 | 100.0 | 56 |  |
| Nicorandil (C01DX16) | 0.0041 | 0.0 | 100.0 | 57 |  |
| Phenylephrine (C01CA06) | 0.003 | 0.0 | 100.0 | 58 |  |
| Flecainide (C01BC04) | 0.0018 | 0.0 | 100.0 | 59 |  |
| Milrinone (C01CE02) | 0.0009 | 0.0 | 100.0 | 60 |  |
| Labetalol (C07AG01) | 0.0006 | 0.0 | 100.0 | 61 |  |
| Ephedrine (C01CA26) | 0.0001 | 0.0 | 100.0 | 62 |  |
| Indapamide (C03BA11) | 0 | 0.0 | 100.0 | 63 |  |
| Norepinephrine (C01CA03) | 0 | 0.0 | 100.0 | 64 |  |
| **Total** | **705.4607** |  |  |  |  |

**Supplementary Table 3: Average monthly cardiovascular medicine utilisation (DDD per 100 bed-days) across the study period (2017–2022), demonstrating the absence of a recurrent seasonal pattern**

| **Month** | **DDD per 100 bed-days** |
| --- | --- |
| January | 10.15 |
| February | 7.66 |
| March | 8.56 |
| April | 8.43 |
| May | 13.23 |
| June | 16.33 |
| July | 9.84 |
| August | 10.53 |
| September | 15.24 |
| October | 10.82 |
| November | 9.12 |
| December | 8.34 |
